# Testing Frequency Matters | An Evaluation of the Diagnostic Performance of a SARS-CoV-2 Rapid Antigen Test in United States Correctional Facilities

**DOI:** 10.1101/2022.03.03.22271803

**Authors:** Margaret L. Lind, Olivia L. Schultes, Alexander J. Robertson, Amy J. Houde, Derek A.T. Cummings, Albert I. Ko, Byron S. Kennedy, Robert P. Richeson

## Abstract

**Background:** The CDC recommends serial rapid antigen assay collection within congregate facilities for screening and outbreak testing. Though modeling and observational studies from community and long-term care facilities have shown serial collection provides adequate sensitivity and specificity, the diagnostic accuracy of this testing strategy within correctional facilities remains unknown.

**Methods:** Using Connecticut Department of Corrections (DOC) data from November 21^st^ 2020 to June 15^th^ 2021, we estimated the accuracy of a rapid assay, BinaxNOW, under three collection strategies, a single test in isolation and two and three serial tests separated by 1-4 day intervals. Diagnostic accuracy metrics were estimated in relation to RT-PCRs collected within one day before the first or after the last included rapid antigen tests in a series.

**Results:** Of the 17,669 residents who contributed at least one RT-PCR or rapid antigen during the study period, 3,979 contributed ≥1 paired rapid antigen test series. In relation to RT-PCR, the three-rapid antigen test strategy had a sensitivity of 89.6% (95% confidence intervals: 86.1-92.6%) and specificity of 97.2% (CI: 95.1-98.3%). The sensitivities for two and one-rapid antigen test strategy were 75.2% and 52.8%, respectively, and the specificities were 98.5% and 99.4%, respectively. The sensitivity was higher among symptomatic residents and when the RT-PCR was collected before the rapid antigen tests.

**Conclusions:** We found the serial collection of an antigen test resulted in high diagnostic accuracy. These findings support serial testing within correctional facilities for outbreak investigation, screening, and when rapid detection is required (such as intakes or transfers).

## Introduction

Within the United States, state and federal run correctional facilities have experienced high transmission rates of SARS-CoV-2 and remain high-risk settings for COVID-19.[1,2] In fact, data from September through November 2020 show that residents of Federal Bureau of Prisons were 4.7 times more likely to become infected with SARS-CoV-2 and 2.6 times more likely to die from COVID-19 than general US residents.[2] Despite the development of COVID-19 vaccines and vaccination programs for incarcerated populations, vaccine coverage remains below that needed for population level protection.[3–10] Rapid and accurate SAR-CoV-2 testing will therefore remain a key component of infection prevention within correctional facilities.

In August 2020, the Food and Drug Administration (FDA) issued an emergency use authorization of Abbott’s BinaxNOW, a COVID-19 rapid antigen test.[11,12] Compared with reverse transcription polymerase chain reaction (RT-PCR), the rapid turnaround time and low cost of rapid antigen tests makes them a cost-effective strategy for congregate settings, where transmission risk is high and implementation of serial mass screening is feasible.[13] Unfortunately, prior studies from community, educational, and long-term care facility settings found the sensitivity of rapid antigen tests to be poor to moderate (53-77%) and to be lower among asymptomatic individuals and individuals early or late in their course of infection.[14–19] These findings, thus, call into question the use of rapid antigen tests as single point of care tests.

Single test collection is not, however, the intended testing strategy for rapid antigen tests outside of healthcare settings.[12,20,21] Instead, both the manufactures and the FDA advice serial collection of at least two tests.[12,20] In alignment with the intended use, the Centers for Disease Control (CDC) recommends serial collection of rapid antigen testing within congregate settings for screening and during outbreaks regardless of symptom presentation.[22] This guidance is supported by modeled evidence that outbreak control depends largely on frequency and speed of testing and observational data showing that serial testing improves the sensitivity of rapid antigen tests in both nursing home and community settings.[13,15,23,24] However, the value of serial testing within correctional facilities remains unknown.

Herein, we present findings of a study that evaluated the accuracy of serial BinaxNOW rapid antigen testing during a mass screening and testing program implemented by the Connecticut Department of Corrections (DOC) during the COVID-19 pandemic wave in late 2020 and early 2021. We specifically compared the accuracy of the rapid antigen test relative to RT-PCR under three collection strategies: serial testing of up to three negative rapid antigen tests, serial testing of up to two negative rapid antigen tests, and rapid antigen collection in isolation.

## Methods

### Setting and Specimen Collection

On November 21, 2020, the DOC initiated the implementation of rapid antigen test (BinaxNOW) collection into their SARS-CoV-2 testing program for symptomatic testing, contact tracing, testing of residents undergoing admission to the correctional facilities and inter-facility transfer and mass voluntary asymptomatic screening. For each instance of rapid antigen use, the DOC guidelines recommend the serial collection of up to three negative rapid antigen tests taken on day one, four, and seven. Due to concerns around the sensitivity of the rapid antigen test, phased implementation of rapid antigen testing with confirmatory RT-PCR was performed. While undergoing serial test collection, residents of DOC facilities were placed in quarantine or isolation.

Trained medical staff collected anterior nasal and nasopharyngeal swab specimens for rapid antigen and RT-PCR testing. Quest Diagnostic facilities performed SARS-CoV-2 RT-PCR testing of nasopharnyngeal swab specimens and defined positive tests as having a cycle threshold value less than 40.[25] At the time rapid antigen test collection was implemented, the DOC oversaw 17 facilities with a resident census of around 10,000 residents (9,945 residents on July 1^st^, 2020).[26,27] The Yale University Institutional Review Board classified this study as public health surveillance.

### Data Collection and Cohort Development

Using resident and testing data queried from internally maintained DOC databases, we retrospectively identified all rapid antigen (BinaxNOW) and RT-PCR testing records from November 21^st^, 2020 to June 15^th^, 2021. Following the removal of duplicated records (same day, assay, and assay results) and reporting errors (negative recorded on the same day as a positive), we identified rapid antigen test series (rapid antigen tests collected within one and four days of each other or tests collected in the absence of any test in the prior or following four days). We then retained the first three test of each series, resulting in series of between one and three rapid antigen tests (eFigure1).

We paired the rapid antigen test series with RT-PCR tests collected between one day prior to the first and one day following the last rapid antigen test of the series (eFigure1). In the event of multiple RT-PCR matches per series, ordered preference was given to positive RT-PCRs, RT-PCRs collected on the same day or between rapid antigen tests of a series, and RT-PCRs collected before the rapid antigen test series. We defined symptoms as the presence or absence of COVID-19 related symptoms reported at the time of the rapid antigen test.[28]

### Statistical Approach: Diagnostic Accuracy

The characteristics of residents with at least one RT-PCR paired rapid antigen test series were summarized by presence of RT-PCR positive SARS-CoV-2 events using counts and percentages for categorical factors and means and standard errors for continuous factors. We determined the diagnostic accuracy of rapid antigen test in relation to RT-PCR using sensitivity, specificity, and positive and negative predictive value (PPV/NPV). We estimated the sensitivity and specificity of the first, second, and third test of all series using generalized estimating equations (GEE) with robust standard errors and a logit link (eFigure2). With these estimates, we calculated the sensitivity and specificity for each testing strategy using the following equations[29]:

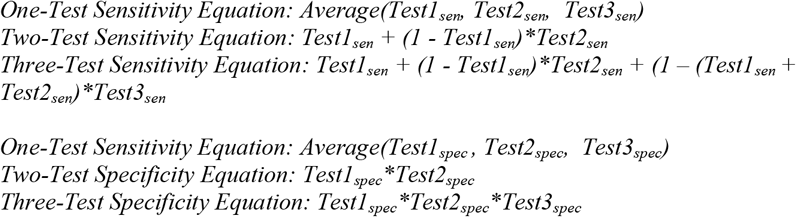

Where Test1_sen/spec_ was the sensitivity or specificity of the first rapid antigen test of all series, Test2_sen/spec_ was the sensitivity or specificity of the second rapid antigen test of all series of two or more rapid antigen tests, and Test3_sen/spec_ was the sensitivity or specificity of the third rapid antigen test of all series of three rapid antigen tests (eFigure2). We propagated the uncertainty through the serial testing equations using posterior simulation of 1000 random draws of the GEE estimate.[30,31] Additionally, we estimated the diagnostic accuracy stratified by the following *a priori* selected factors: age, symptom presence, and test order. We tested for additive differences in the diagnostic accuracy of the stratified samples by subtracting the draws of each sample from a selected reference category.

For the PPV and NPV, we simulated 1000 average daily prevalence estimates using a Poisson regression with an outcome of positive SARS-CoV-2 test (either rapid antigen or RT-PCR test). To reduce the risk of including multiple positive tests from the same testing event, we excluded positive events within 5 days of each other. With the estimated prevalence, sensitivity, and specificity, we estimated the PPV and NPV at the draw level.[32] For each accuracy metric, we calculated 95% confidence intervals as the 2.5 and 97.5 percentiles.[31] All analyses were conducted in R 4.1.0 using the geepack and multcomp packages.[33,34]

### Sensitivity Analyses

Because of the retrospective nature of the presented analysis, we had to make assumptions around test selection and the appropriate timing of RT-PCR and rapid antigen testing among included pairs. To test the robustness of our findings, we performed sensitivity analyses where we selected first, second and third series tests at random instead of the observed order, limited to rapid antigen test series where tests were collected exactly three days apart, and invoked different selection approaches in the event of multiple RT-PCRs linked to a rapid antigen test series. Unless otherwise stated, all sensitivity analyses were performed among the full sample and mirrored the primary analysis for all but their stated variation.

## Results

Between November 21^st^ 2020 and June 15^th^ 2021, 128,986 RT-PCR and 30,112 rapid antigen tests were collected among 17,669 DOC residents (Figure1). Of the 3,367 residents who contributed at least one RT-PCR paired rapid antigen test series, 583 (17.3%) contributed at least one rapid antigen test series paired with a positive RT-PCR. The majority of residents were male (74.9%), and the most frequently observed race was Black (40.7%). Individuals who experienced a paired RT-PCR positive SARS-CoV-2 event were demographically similar to individuals who did not experience a paired RT-PCR positive SARS-CoV-2 event.

**Figure 1:**
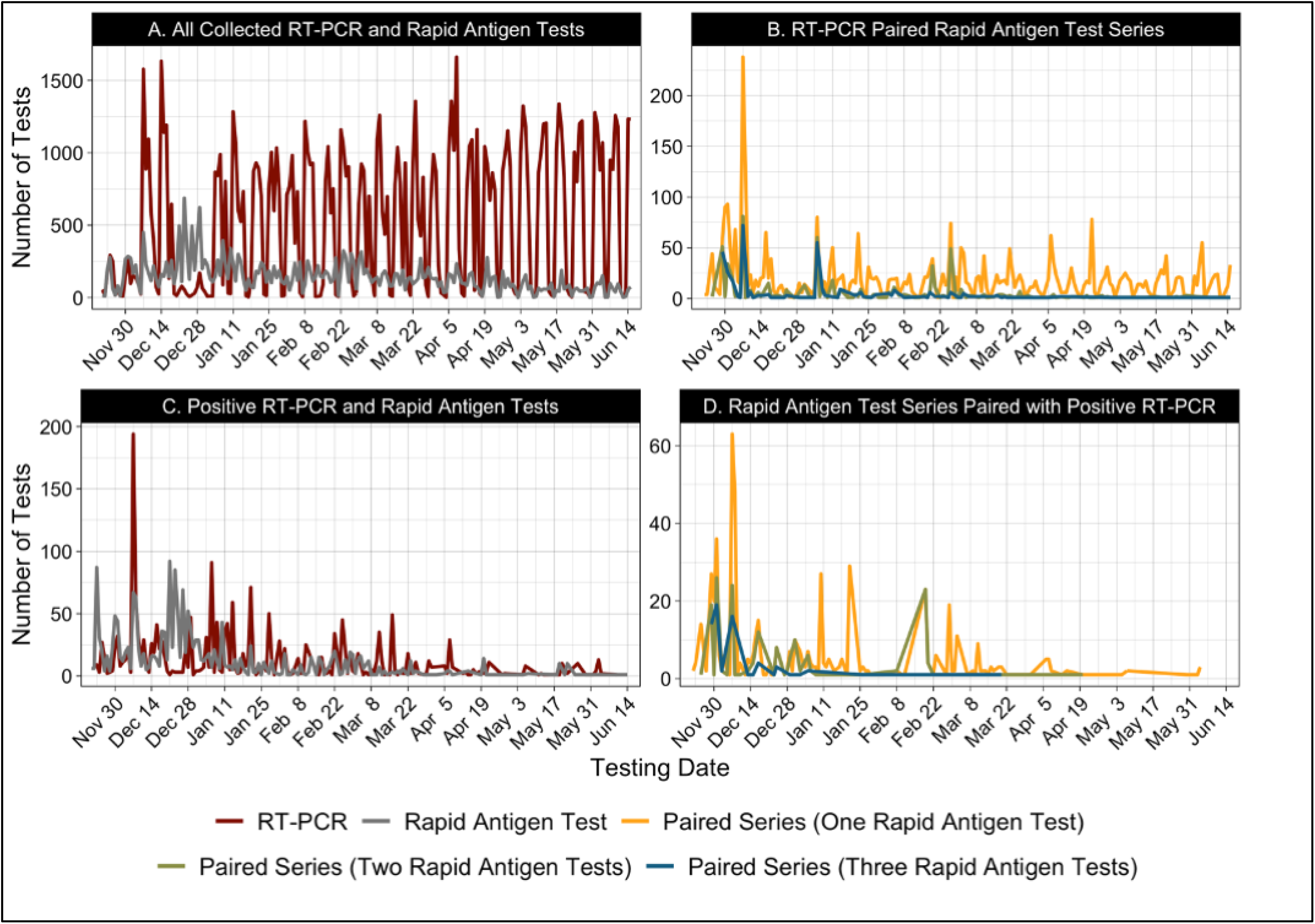
SARS-CoV-2 daily testing volumes from Connecticut run correctional facilities by date for A. all collected paired and unpaired tests, B. RT-PCR paired rapid antigen test series, C. all positive RT-PCR and rapid antigen tests, and D. positive RT-PCR paired rapid antigen test series. Red, Grey, Yellow, Green, Blue lines show volumes of SARS-CoV-2 RT-PCR only, rapid antigen test only, paired RT-PCR series of one rapid antigen test, paired RT-PCR series of two rapid antigen tests, paired RT-PCR series of three rapid antigen tests, respectively. Series were defined as rapid antigen tests collected within one and four days of each other or tests collected in the absence of any test in the prior or following four days. Paired series were defined as rapid antigen test series with RT-PCR collected between one day prior to the first and one day following the last test of the series.

Residents contributed an average of 1.2 RT-PCR paired rapid antigen test series (3,884 pairs/3,367 residents) and six people contributed more than one positive RT-PCR paired rapid antigen test series (Table1). Among the 3,884 included series, 692 consisted of at least two rapid antigen tests and 358 consisted of three rapid antigen tests (Figure 2). The second rapid antigen test of included series were more likely to be collected prior to the paired RT-PCR (62.1%) than the first (37.3%) and third (23.7%) test of included series. The proportion of tests collected among symptomatic and asymptomatic individuals was similar between the first, second, and third tests of included series (eTable1).

**Table1:**
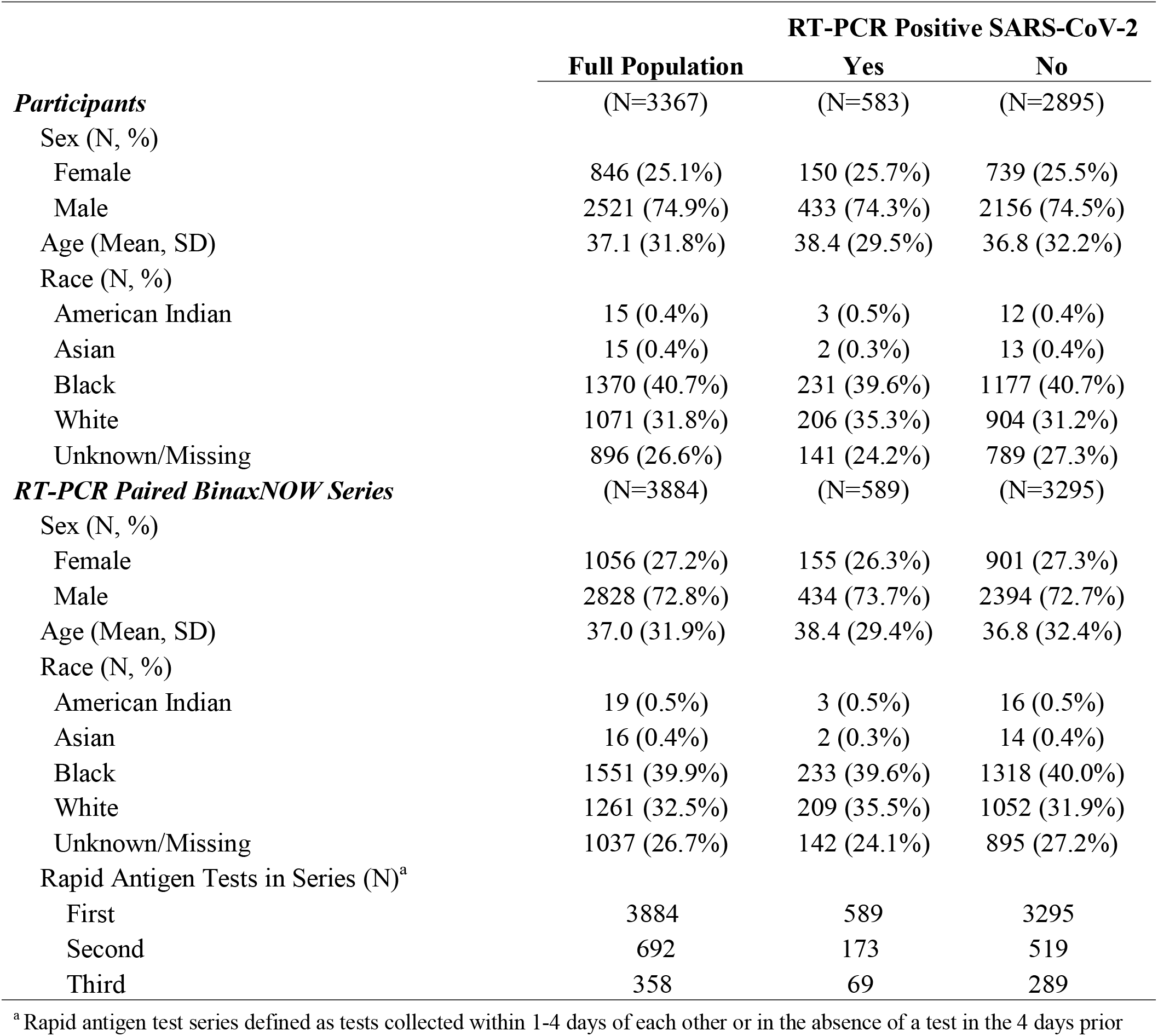
**Demographic Characteristics of Residents of Connecticut State Correctional Facilities with Time Matched BinaxNOW and RT-PCR tests by Occurrence of SARS-CoV-2 Infection**

|  | Full Population | RT-PCR Positive SARS-CoV-2 |  |
| --- | --- | --- | --- |
|  |  | Yes | No |
| *Participants* | (N=3367) | (N=583) | (N=2895) |
| Sex (N, %) |  |  |  |
| Female | 846 (25.1%) | 150 (25.7%) | 739 (25.5%) |
| Male | 2521 (74.9%) | 433 (74.3%) | 2156 (74.5%) |
| Age (Mean, SD) | 37.1 (31.8%) | 38.4 (29.5%) | 36.8 (32.2%) |
| Race (N, %) |  |  |  |
| American Indian | 15 (0.4%) | 3 (0.5%) | 12 (0.4%) |
| Asian | 15 (0.4%) | 2 (0.3%) | 13 (0.4%) |
| Black | 1370 (40.7%) | 231 (39.6%) | 1177 (40.7%) |
| White | 1071 (31.8%) | 206 (35.3%) | 904 (31.2%) |
| Unknown/Missing | 896 (26.6%) | 141 (24.2%) | 789 (27.3%) |
| *RT-PCR Paired BinaxNOW Series* | (N=3884) | (N=589) | (N=3295) |
| Sex (N, %) |  |  |  |
| Female | 1056 (27.2%) | 155 (26.3%) | 901 (27.3%) |
| Male | 2828 (72.8%) | 434 (73.7%) | 2394 (72.7%) |
| Age (Mean, SD) | 37.0 (31.9%) | 38.4 (29.4%) | 36.8 (32.4%) |
| Race (N, %) |  |  |  |
| American Indian | 19 (0.5%) | 3 (0.5%) | 16 (0.5%) |
| Asian | 16 (0.4%) | 2 (0.3%) | 14 (0.4%) |
| Black | 1551 (39.9%) | 233 (39.6%) | 1318 (40.0%) |
| White | 1261 (32.5%) | 209 (35.5%) | 1052 (31.9%) |
| Unknown/Missing | 1037 (26.7%) | 142 (24.1%) | 895 (27.2%) |
| Rapid Antigen Tests in Series (N)<sup>a</sup> |  |  |  |
| First | 3884 | 589 | 3295 |
| Second | 692 | 173 | 519 |
| Third | 358 | 69 | 289 |

<sup>a</sup> Rapid antigen test series defined as tests collected within 1-4 days of each other or in the absence of a test in the 4 days prior

**Figure 2:**
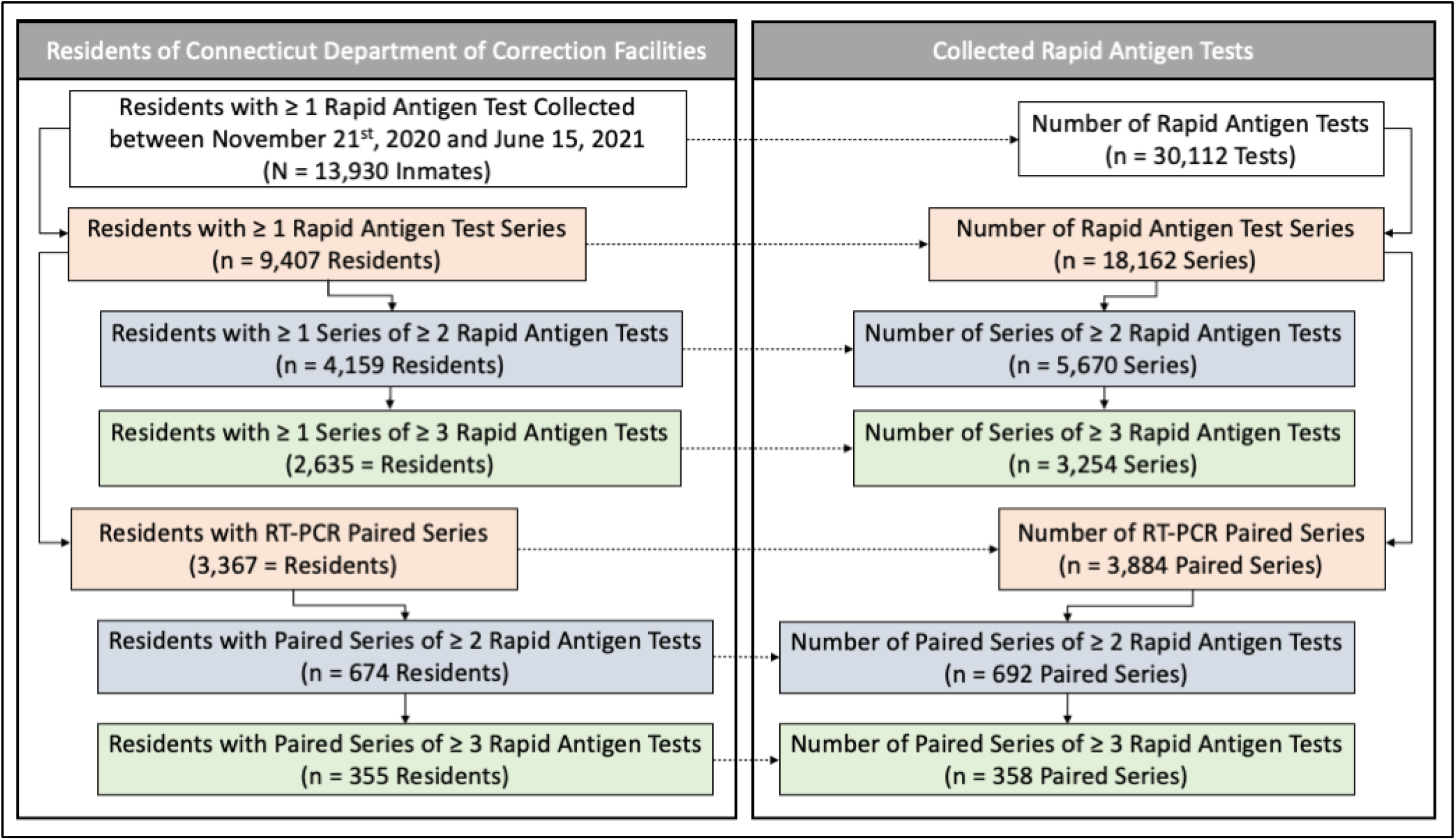
Flowchart of residents of Connecticut run correctional facilities and rapid antigen test series included in our analysis. Series were defined as rapid antigen tests collected within one and four days of each other or tests collected in the absence of any test in the prior or following four days. Paired series were defined as rapid antigen test series with RT-PCR collected between one day prior to the first and one day following the last test of the series.

Most rapid antigen test of included series were negative (first test: 91.6%, second test: 86.7%, third test: 88.5%). Of the negative rapid antigen tests, most were paired with negative RT-PCRs (first test: 84.3%; second test: 73.8%; third test: 80.4%; eTable2). Relative to RT-PCR, the sensitivity of the first, second and third-rapid antigen tests of included series were 51.6% (95% confidence interval: 47.6-55.6%), 48.6% (CI: 41.2-56.0%) and 58.0% (CI: 46.2-69.2%), respectively. This resulted in sensitivities of 75.2% (CI: 71.0-79.2%) and 89.6% (86.1-92.6%) for the serial collection of two and three rapid antigen tests, respectively. The specificity of each rapid antigen test of included series was above 98%. The specificity of two and three serially collected rapid antigen tests was 97.8% (96.4-98.6%) and 97.2% (95.1-98.4%), respectively (Table2). The PPV, based on an observed prevalence of 57 (CI: 53-62) cases per 100,000 residents, was highest for the one-rapid antigen test strategy (63.5%) and lowest for the three-rapid antigen test strategies (50.1%). The NPV for each rapid antigen test collection strategy was high (one-rapid antigen test: 98.6%, two-rapid antigen tests: 99.2%, three-rapid antigen tests: 99.7%, eTable3).

**Table 2:**
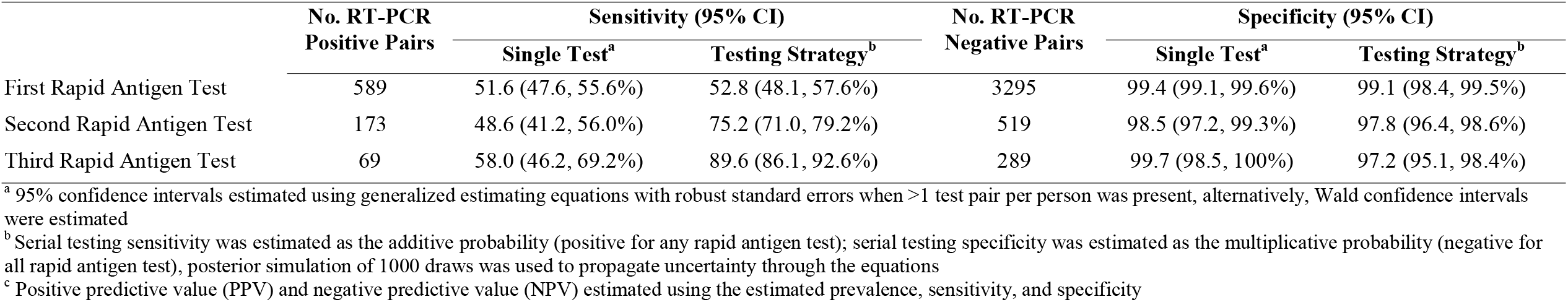
**Rapid Antigen Test Accuracy Relative to RT-PCR Among Residents of Connecticut State Correctional Facilities Under Varying Collection Strategies**

The sensitivity for each collection strategy was significantly higher among symptomatic residents (one-rapid antigen test: 62.9%, two-rapid antigen tests: 82.2%, three-rapid antigen tests: 95.6%) than among asymptomatic patients (one-rapid antigen test: 47.4%, two-rapid antigen tests: 72.0%, three-rapid antigen tests: 85.5%). The sensitivity was higher when the rapid antigen test was collected on the same day as the RT-PCR (one-rapid antigen test: 73.5%; two-rapid antigen tests: 93.6%, three-rapid antigen tests: no sample) than when the rapid antigen test was collected prior (one-rapid antigen test: 15.2%; two-rapid antigen tests: 26.9%, three-rapid antigen tests: 39.4%). The sensitivity of the three-rapid antigen test collection strategy was significantly lower for individuals more than 37 years of age than individuals 37 or fewer years of age (difference: 8.3 CI: 1.8-14.6; Table3).

**Table 3:**
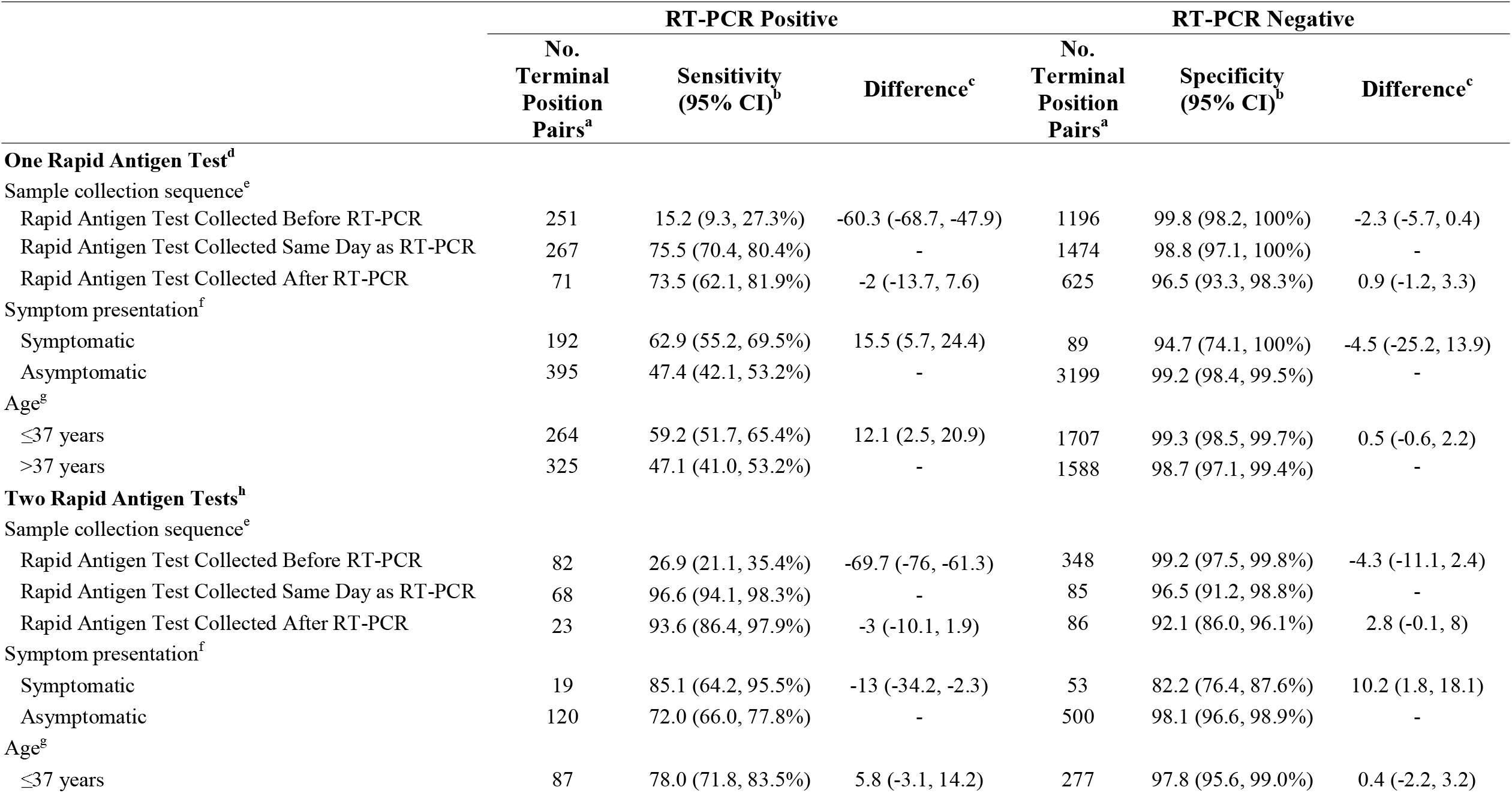
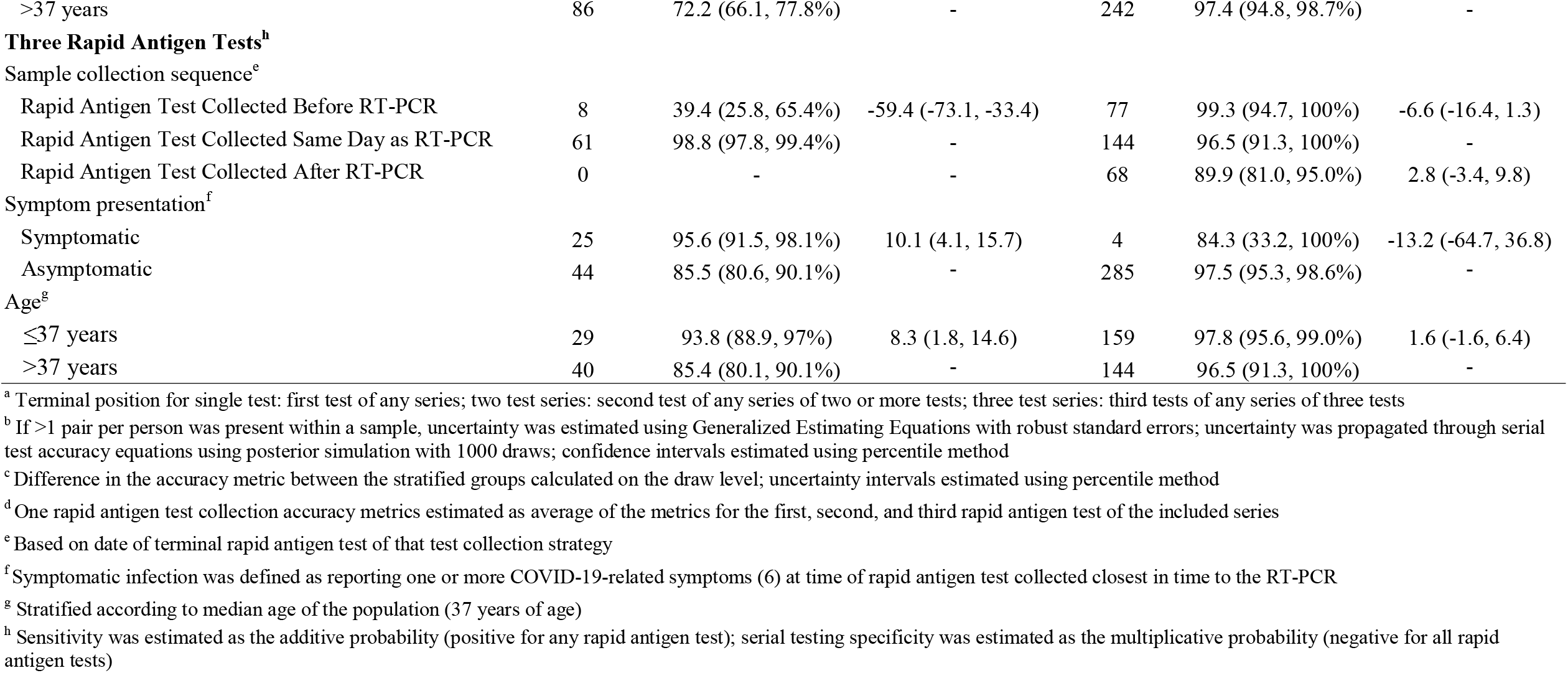
**Rapid Antigen Test Accuracy Relative to RT-PCR Among Residents of Connecticut State Correctional Facilities Under Varying Collection Strategies by Demographic and Clinical Characteristics**

|  | RT-PCR Positive |  |  | RT-PCR Negative |  |  |
| --- | --- | --- | --- | --- | --- | --- |
|  | No.<br>Terminal<br>Position<br>Pairs<sup>a</sup> | Sensitivity<br>(95% CI)<sup>b</sup> | Difference<sup>c</sup> | No.<br>Terminal<br>Position<br>Pairs<sup>a</sup> | Specificity<br>(95% CI)<sup>b</sup> | Difference<sup>c</sup> |
| **One Rapid Antigen Test<sup>d</sup>** |  |  |  |  |  |  |
| Sample collection sequence<sup>e</sup> |  |  |  |  |  |  |
| Rapid Antigen Test Collected Before RT-PCR | 251 | 15.2 (9.3, 27.3%) | -60.3 (-68.7, -47.9) | 1196 | 99.8 (98.2, 100%) | -2.3 (-5.7, 0.4) |
| Rapid Antigen Test Collected Same Day as RT-PCR | 267 | 75.5 (70.4, 80.4%) | - | 1474 | 98.8 (97.1, 100%) | - |
| Rapid Antigen Test Collected After RT-PCR | 71 | 73.5 (62.1, 81.9%) | -2 (-13.7, 7.6) | 625 | 96.5 (93.3, 98.3%) | 0.9 (-1.2, 3.3) |
| Symptom presentation<sup>f</sup> |  |  |  |  |  |  |
| Symptomatic | 192 | 62.9 (55.2, 69.5%) | 15.5 (5.7, 24.4) | 89 | 94.7 (74.1, 100%) | -4.5 (-25.2, 13.9) |
| Asymptomatic | 395 | 47.4 (42.1, 53.2%) | - | 3199 | 99.2 (98.4, 99.5%) | - |
| Age<sup>g</sup> |  |  |  |  |  |  |
| ≤37 years | 264 | 59.2 (51.7, 65.4%) | 12.1 (2.5, 20.9) | 1707 | 99.3 (98.5, 99.7%) | 0.5 (-0.6, 2.2) |
| >37 years | 325 | 47.1 (41.0, 53.2%) | - | 1588 | 98.7 (97.1, 99.4%) | - |
| **Two Rapid Antigen Tests<sup>h</sup>** |  |  |  |  |  |  |
| Sample collection sequence<sup>e</sup> |  |  |  |  |  |  |
| Rapid Antigen Test Collected Before RT-PCR | 82 | 26.9 (21.1, 35.4%) | -69.7 (-76, -61.3) | 348 | 99.2 (97.5, 99.8%) | -4.3 (-11.1, 2.4) |
| Rapid Antigen Test Collected Same Day as RT-PCR | 68 | 96.6 (94.1, 98.3%) | - | 85 | 96.5 (91.2, 98.8%) | - |
| Rapid Antigen Test Collected After RT-PCR | 23 | 93.6 (86.4, 97.9%) | -3 (-10.1, 1.9) | 86 | 92.1 (86.0, 96.1%) | 2.8 (-0.1, 8) |
| Symptom presentation<sup>f</sup> |  |  |  |  |  |  |
| Symptomatic | 19 | 85.1 (64.2, 95.5%) | -13 (-34.2, -2.3) | 53 | 82.2 (76.4, 87.6%) | 10.2 (1.8, 18.1) |
| Asymptomatic | 120 | 72.0 (66.0, 77.8%) | - | 500 | 98.1 (96.6, 98.9%) | - |
| Age<sup>g</sup> |  |  |  |  |  |  |
| ≤37 years | 87 | 78.0 (71.8, 83.5%) | 5.8 (-3.1, 14.2) | 277 | 97.8 (95.6, 99.0%) | 0.4 (-2.2, 3.2) |
| >37 years | 86 | 72.2 (66.1, 77.8%) | - | 242 | 97.4 (94.8, 98.7%) | - |
| **Three Rapid Antigen Tests<sup>h</sup>** |  |  |  |  |  |  |
| Sample collection sequence<sup>e</sup> |  |  |  |  |  |  |
| Rapid Antigen Test Collected Before RT-PCR | 8 | 39.4 (25.8, 65.4%) | -59.4 (-73.1, -33.4) | 77 | 99.3 (94.7, 100%) | -6.6 (-16.4, 1.3) |
| Rapid Antigen Test Collected Same Day as RT-PCR | 61 | 98.8 (97.8, 99.4%) | - | 144 | 96.5 (91.3, 100%) | - |
| Rapid Antigen Test Collected After RT-PCR | 0 | - | - | 68 | 89.9 (81.0, 95.0%) | 2.8 (-3.4, 9.8) |
| Symptom presentation<sup>f</sup> |  |  |  |  |  |  |
| Symptomatic | 25 | 95.6 (91.5, 98.1%) | 10.1 (4.1, 15.7) | 4 | 84.3 (33.2, 100%) | -13.2 (-64.7, 36.8) |
| Asymptomatic | 44 | 85.5 (80.6, 90.1%) | - | 285 | 97.5 (95.3, 98.6%) | - |
| Age<sup>g</sup> |  |  |  |  |  |  |
| ≤37 years | 29 | 93.8 (88.9, 97%) | 8.3 (1.8, 14.6) | 159 | 97.8 (95.6, 99.0%) | 1.6 (-1.6, 6.4) |
| >37 years | 40 | 85.4 (80.1, 90.1%) | - | 144 | 96.5 (91.3, 100%) | - |

<sup>a</sup> Terminal position for single test: first test of any series; two test series: second test of any series of two or more tests; three test series: third tests of any series of three tests
<sup>b</sup> If >1 pair per person was present within a sample, uncertainty was estimated using Generalized Estimating Equations with robust standard errors; uncertainty was propagated through serial test accuracy equations using posterior simulation with 1000 draws; confidence intervals estimated using percentile method
<sup>c</sup> Difference in the accuracy metric between the stratified groups calculated on the draw level; uncertainty intervals estimated using percentile method
<sup>d</sup> One rapid antigen test collection accuracy metrics estimated as average of the metrics for the first, second, and third rapid antigen test of the included series
<sup>e</sup> Based on date of terminal rapid antigen test of that test collection strategy
<sup>f</sup> Symptomatic infection was defined as reporting one or more COVID-19-related symptoms (6) at time of rapid antigen test collected closest in time to the RT-PCR
<sup>g</sup> Stratified according to median age of the population (37 years of age)
<sup>h</sup> Sensitivity was estimated as the additive probability (positive for any rapid antigen test); serial testing specificity was estimated as the multiplicative probability (negative for all rapid antigen tests)

The specificity for each rapid antigen test collection strategy was significantly lower among symptomatic residents (one-rapid antigen test: 94.7%, two-rapid antigen tests: 85.1%, three-rapid antigen tests: 84.3%) than asymptomatic residents (one-rapid antigen test: 99.2%, two-rapid antigen tests: 98.1%, three-rapid antigen tests: 97.5%). The specificities did not vary significantly by age or timing of tests (Table3).

### Sensitivity Analyses

We performed three sets of sensitivity analyses to test the robustness of our findings to varying rapid antigen test collection orders, rapid antigen test collection windows, and RT-PCR pair selection. The sensitivity of the one-rapid antigen test, two-rapid antigen test, and three-rapid antigen test strategies ranged from 51.0-75.4%, 76.3-86.1%, 87.2-100%, respectively. The highest sensitivities were observed when we restricted to rapid antigen test series with tests collected exactly three days apart (eTable4). The lowest sensitivities were observed when we randomly selected the first, second, and third test of each series (eTable5). As with the primary analysis, we observed high specificities for each testing strategy (one-rapid antigen test: 99.0-99.6%, two-rapid antigen test: 97.7-98.9%, three-rapid antigen test: 96.9-98.9%; eTables4-5).

## Discussion

We evaluated the predictive accuracy of the rapid COVID-19 antigen test, BinaxNOW, in relation to RT-PCR under three different collection strategies (a single test in isolation and two and three serial tests separated by 1-4 day intervals) among residents of Connecticut state prisons and jails between November 21^st^, 2020 and June 15^th^, 2021. The three test strategy is currently recommended by Connecticut DOC.[35] In alignment with diagnostic accuracy estimates from other congregate settings, we found that rapid antigen tests had a low sensitivity when collected in isolation, but the sensitivity increased significantly when rapid antigen tests were collected in pairs and triplets.[14,15,23] Further, we found that the sensitivity for all rapid antigen test collection strategies was significantly higher among symptomatic individuals than asymptomatic individuals.

In relation to RT-PCR, we found the current DOC rapid antigen test collection strategy, three-rapid antigen test, had a high sensitivity (90%) and specificity (97%). While the specificity of the two and one-rapid antigen test collection strategies were higher (98% and 99%, respectively), the sensitivity for these less intensive collection strategies were significantly lower (75% and 53%, respectively). These findings suggest that, among 100 residents infected with SARS-CoV-2, 90 would be captured by the three-rapid antigen test strategy. Compared with this strategy, the two-rapid antigen test and the one-rapid antigen test strategies would miss an additional 15 and 37 infected individuals, respectively. Conversely, the three-rapid antigen test strategy would only misdiagnose three out of 100 uninfected residents. Though this is two more than the one-rapid antigen test strategy, the cost of false negatives (missed infections) far outweighs the cost of false positives (excess isolation) under scenarios of highly transmissible infectious diseases, such as SARS-CoV-2.[36]

In alignment with prior studies, we found the sensitivity of rapid antigen testing to be significantly higher later in the course of infection (rapid antigen tests collected on the same day or after RT-PCR) and among residents exhibiting COVID-19 symptoms.[37,38] However, contrary to previous findings, we found the specificity to be significantly lower among symptomatic residents than among asymptomatic residents.[38–40] While, the magnitude of this difference was minimal for the one-rapid antigen test strategy (specificity among symptomatic residents was 5% lower than asymptomatic residents), the difference was larger (13%) for the three-rapid antigen test strategy. Despite this meaningful difference in the specificity among symptomatic and asymptomatic residents under the three-rapid antigen test strategy, the specificity among symptomatic residents remained high (84%). Thus, this difference in specificity does not invalidate the use of the three-test strategy for symptomatic residents of correctional facilities.

Taken collectively, our findings support the use of serial rapid antigen testing under scenarios when PCR turnround time is long, rapid detection is required, or when isolating/quarantining all exposed individuals is unfeasible. Such scenarios include times of unknown exposure (intake and transfers), contact tracing, asymptomatic and symptomatic screening, and during outbreak investigations. These recommendations stem from the rapid turnaround time and low cost of a single test, the collectively high diagnostic accuracy of serial collection, and the ability of serial collection strategies to detect events under continuous exposure scenarios. Though our findings speak predominately to the value of serial collection resulting from a single exposure, serial collection provides additional benefit through capture of infections from exposures that occurred immediately prior to or following the first collected test.[13] The combination of these benefits, thus, may results in more rapid isolation of infected individuals, and in turn, a reduction in facility wide transmission.

### Limitations and Strengths

Our study was subject to limitations typical of retrospective diagnostic validation analyses. First, race/ethnicity was missing for a large portion of the population and we were unable to test for differences in test accuracy by race. However, it is unlikely that race would impact the diagnostic accuracy of the rapid antigen test. Second, we relied on a reference outcome of RT-PCR positivity, which is an imperfect indicator of infection. In addition, in the absence of cycle threshold values, we were unable to tests the impact of viral load on rapid antigen test performance. Third, our accuracy estimates relied on collected test that may be biased towards department specific testing practices. However, we observed similar results when we selected the first, second, and third test of each series at random (eTable5). Finally, our study was conducted prior to the large Delta and Omicron waves of 2021. While the diagnostic accuracy of rapid antigen tests likely varies between the different variants, we believe the benefits of serial collection will hold.

Our study had several strengths including our large sample of paired assays collected among a diverse population of Connecticut State correctional facility residents. This large sample allowed us to estimate and compare the diagnostic accuracy of three different collection strategies, including the three-test strategy employed by the Connecticut DOC. Our large sample also allowed us to examine characteristics associated with the accuracy of the rapid antigen test within correctional facilities settings and speak to the use of different collection strategies based on these characteristics. Additionally, through the inclusion of numerous sensitivity analyses, we were able to show that our findings were not the result of the data cleaning or modeling assumptions we employed within this analysis. Finally, we were able to include all unique rapid antigen test sets and account for within person correlation in our uncertainty intervals using generalized estimating equations.

## Conclusion

Compared with singularly collected tests, we found that serial collection of BinaxNOW rapid antigen tests resulted in meaningfully higher sensitivities and comparably high specificities among residents in state correctional facilities. We found this held for both asymptomatic and symptomatic residents and regardless of rapid antigen test collection time relative to RT-PCR collection. These findings speak to the utility of serially collected rapid antigen tests within correctional facilities for asymptomatic and symptomatic screening, contact tracing, and during outbreak investigations. If employed under such scenarios, rapid antigen testing may result in faster isolation of infected individuals and reduce transmission within facilities.

## Data Availability

Data Use Agreement: The data used in this study belongs to the Connecticut Department of Corrections. Qualified researchers may submit a data share request for de-identified patient level data by contacting the corresponding author with a detailed description of the research question.

## Additional Contributions

We thank the Connecticut Department of Corrections and the healthcare workers in each correctional facility for administrative support and administering RT-PCR and rapid antigen tests. None of the individuals received compensation for their contribution.

## Data Use Agreement

The data used in this study belongs to the Connecticut Department of Corrections. Qualified researchers may submit a data share request for de-identified patient level data by contacting the corresponding author with a detailed description of the research question.

## Author Contributions

Margaret L. Lind had full access to all the data in the study and takes responsibility for the integrity of the data and the accuracy of the data analysis.

*Concept and design:* Ko, Kennedy, Richeson

*Acquisition, analysis, or interpretation of data:* Schultes, Lind

*Drafting of the manuscript:* Lind, Schultes, Robertson, Ko, Cummings

*Critical revision of the manuscript for important intellectual content:* All authors.

*Statistical analysis:* Lind

*Administrative, technical, or material support:*

*Supervision:* Ko, Kennedy, Richeson, Cummings

## References

1. Hagan LM, McCormick DW, Lee C, et al. Outbreak of SARS-CoV-2 B.1.617.2 (Delta) Variant Infections Among Incarcerated Persons in a Federal Prison - Texas, July-August 2021. MMWR Morb Mortal Wkly Rep 2021; 70:1349–1354.

2. Toblin RL, Hagan LM. COVID-19 Case and Mortality Rates in the Federal Bureau of Prisons. Am J Prev Med 2021; 61:120–123.

3. Chin ET, Leidner D, Ryckman T, et al. Covid-19 Vaccine Acceptance in California State Prisons. New England Journal of Medicine 2021; 385:374–376.

4. Chemaitelly H, Tang P, Hasan MR, et al. Waning of BNT162b2 Vaccine Protection against SARS-CoV-2 Infection in Qatar. New England Journal of Medicine 2021; Available at: https://www.nejm.org/doi/10.1056/NEJMoa2114114. Accessed 7 October 2021.

5. Goldberg Y, Mandel M, Bar-On YM, et al. Waning Immunity after the BNT162b2 Vaccine in Israel. New England Journal of Medicine 2021; Available at: https://www.nejm.org/doi/10.1056/NEJMoa2114228. Accessed 29 October 2021.

6. Garcia-Beltran WF, Lam EC, St. Denis K, et al. Multiple SARS-CoV-2 variants escape neutralization by vaccine-induced humoral immunity. medRxiv 2021; :2021.02.14.21251704.

7. Tartof SY. Effectiveness of mRNA BNT162b2 COVID-19 vaccine up to 6 months in a large integrated health system in the USA: a retrospective cohort study. :10.

8. Fowlkes A. Effectiveness of COVID-19 Vaccines in Preventing SARS-CoV-2 Infection Among Frontline Workers Before and During B.1.617.2 (Delta) Variant Predominance — Eight U.S. Locations, December 2020–August 2021. MMWR Morb Mortal Wkly Rep 2021; 70. Available at: https://www.cdc.gov/mmwr/volumes/70/wr/mm7034e4.htm. Accessed 7 October 2021.

9. Kustin T, Harel N, Finkel U, et al. Evidence for increased breakthrough rates of SARS-CoV-2 variants of concern in BNT162b2-mRNA-vaccinated individuals. Nat Med 2021; 27:1379–1384.

10. Hagan LM, Dusseau C, Crockett M, Rodriguez T, Long MJ. COVID-19 vaccination in the Federal Bureau of Prisons, December 2020—April 2021. Vaccine 2021; 39:5883–5890.

11. FDA Media. COVID-19 Update: FDA Authorizes First Diagnostic Test Where Results Can Be Read Directly From Testing Card. FDA, 2020. Available at: https://www.fda.gov/news-events/press-announcements/covid-19-update-fda-authorizes-first-diagnostic-test-where-results-can-be-read-directly-testing-card. Accessed 12 August 2021.

12. Abbott. Abbott’s BinaxNOWTM Rapid Antigen Self Test Receives FDA Emergency Use Authorization for Asymptomatic, Over-the-Counter, Non-Prescription, Multi-Test Use. Available at: https://abbott.mediaroom.com/2021-03-31-Abbotts-BinaxNOW-TM-Rapid-Antigen-Self-Test-Receives-FDA-Emergency-Use-Authorization-for-Asymptomatic-Over-the-Counter-Non-Prescription-Multi-Test-Use. Accessed 8 November 2021.

13. Larremore DB, Wilder B, Lester E, et al. Test sensitivity is secondary to frequency and turnaround time for COVID-19 screening. Sci Adv 2021; 7:eabd5393.

14. Okoye NC, Barker AP, Curtis K, et al. Performance Characteristics of BinaxNOW COVID-19 Antigen Card for Screening Asymptomatic Individuals in a University Setting. J Clin Microbiol 2021; 59:e03282–20.

15. McKay SL, Tobolowsky FA, Moritz ED, et al. Performance Evaluation of Serial SARS-CoV-2 Rapid Antigen Testing During a Nursing Home Outbreak. Ann Intern Med 2021; 174:945–951.

16. Sood N, Shetgiri R, Rodriguez A, et al. Evaluation of the Abbott BinaxNOW rapid antigen test for SARS-CoV-2 infection in children: Implications for screening in a school setting. PLoS One 2021; 16:e0249710.

17. Dinnes J, Deeks JJ, Berhane S, et al. Rapid, point_Jof_Jcare antigen and molecular_Jbased tests for diagnosis of SARS_JCoV_J2 infection. Cochrane Database of Systematic Reviews 2021; Available at: https://www.cochranelibrary.com/cdsr/doi/10.1002/14651858.CD013705.pub2/full. Accessed 16 August 2021.

18. Brümmer LE, Katzenschlager S, Gaeddert M, et al. Accuracy of novel antigen rapid diagnostics for SARS-CoV-2: A living systematic review and meta-analysis. PLoS Med 2021; 18:e1003735.

19. Pilarowski G, Lebel P, Sunshine S, et al. Performance Characteristics of a Rapid Severe Acute Respiratory Syndrome Coronavirus 2 Antigen Detection Assay at a Public Plaza Testing Site in San Francisco. J Infect Dis 2021; 223:1139–1144.

20. FDA. Fact Sheet For Individuals | Abbott Diagnostics | BinaxNOW COVID-19 Ag Card. 2022; Available at: chrome-extension://efaidnbmnnnibpcajpcglclefindmkaj/viewer.html?pdfurl=https%3A%2F%2Fhttps://www.fda.gov%2Fmedia%2F141568%2Fdownload&clen=274942&chunk=true. Accessed 18 February 2022.

21. FDA. Fact Sheet For Healthcare Providers | Abbott Diagnostics | BinaxNOW COVID-19 Ag Card. 2022; Available at: chrome-extension://efaidnbmnnnibpcajpcglclefindmkaj/viewer.html?pdfurl=https%3A%2F%2Fwww.fda.gov%2Fmedia%2F141568%2Fdownload&clen=274942&chunk=true. Accessed 18 February 2022.

22. CDC. Interim Guidance for Antigen Testing for SARS-CoV-2. 2022. Available at: https://www.cdc.gov/coronavirus/2019-ncov/lab/resources/antigen-tests-guidelines.html. Accessed 18 February 2022.

23. Smith RL, Gibson LL, Martinez PP, et al. Longitudinal Assessment of Diagnostic Test Performance Over the Course of Acute SARS-CoV-2 Infection. The Journal of Infectious Diseases 2021; Available at: https://doi.org/10.1093/infdis/jiab337. Accessed 12 August 2021.

24. Shah MM, Salvatore PP, Ford L, et al. Performance of Repeat BinaxNOW Severe Acute Respiratory Syndrome Coronavirus 2 Antigen Testing in a Community Setting, Wisconsin, November 2020–December 2020. Clinical Infectious Diseases 2021; 73:S54–S57.

25. Quest Diagnostics. SARS-CoV-2 RNA, Qualitative Real-Time RT-PCR (Test Code 39433). Available at: https://www.fda.gov/media/136231/download. Accessed 29 October 2021.

26. Connecticut State Department of Correction. Total Supervised Population Count. Available at: https://portal.ct.gov/DOC/Report/Total-Supervised-Population-Count. Accessed 18 February 2022.

27. Connecticut State Department of Correction. Facilities. Available at: https://portal.ct.gov/DOC/Miscellaneous/Facilities. Accessed 18 February 2022.

28. CDC. Coronavirus Disease 2019 (COVID-19) – Symptoms. 2021. Available at: https://www.cdc.gov/coronavirus/2019-ncov/symptoms-testing/symptoms.html. Accessed 4 August 2021.

29. Weinstein S, Obuchowski NA, Lieber ML. Clinical Evaluation of Diagnostic Tests. American Journal of Roentgenology 2005; 184:14–19.

30. Genders TSS, Spronk S, Stijnen T, Steyerberg EW, Lesaffre E, Hunink MGM. Methods for calculating sensitivity and specificity of clustered data: a tutorial. Radiology 2012; 265:910–916.

31. Burkart KG, Brauer M, Aravkin AY, et al. Estimating the cause-specific relative risks of non-optimal temperature on daily mortality: a two-part modelling approach applied to the Global Burden of Disease Study. The Lancet 2021; 398:685–697.

32. Tenny S, Hoffman MR. Prevalence. In: StatPearls. Treasure Island (FL): StatPearls Publishing, 2022. Available at: http://www.ncbi.nlm.nih.gov/books/NBK430867/. Accessed 16 February 2022.

33. Højsgaard S, Halekoh U, Yan J. The R Package geepack for Generalized Estimating Equations. Journal of Statistical Software 2006; 15:1–11.

34. Hothorn T, Bretz F, Westfall P. Simultaneous inference in general parametric models. Biom J 2008; 50:346–363.

35. CDC. SARS-CoV-2 Antigen Testing in Long Term Care Facilities Considerations for Use in Nursing Homes and other Long-Term Care Facilitiescare Workers. 2020. Available at: https://www.cdc.gov/coronavirus/2019-ncov/hcp/nursing-homes-antigen-testing.html. Accessed 3 November 2021.

36. Hu B, Guo H, Zhou P, Shi Z-L. Characteristics of SARS-CoV-2 and COVID-19. Nat Rev Microbiol 2021; 19:141–154.

37. Mitchell SL, Orris S, Freeman T, et al. Performance of SARS-CoV-2 antigen testing in symptomatic and asymptomatic adults: a single-center evaluation. BMC Infectious Diseases 2021; 21:1071.

38. Pray IW. Performance of an Antigen-Based Test for Asymptomatic and Symptomatic SARS-CoV-2 Testing at Two University Campuses — Wisconsin, September–October 2020. MMWR Morb Mortal Wkly Rep 2021; 69. Available at: https://www.cdc.gov/mmwr/volumes/69/wr/mm695152a3.htm. Accessed 16 February 2022.

39. Prince-Guerra JL. Evaluation of Abbott BinaxNOW Rapid Antigen Test for SARS-CoV-2 Infection at Two Community-Based Testing Sites — Pima County, Arizona, November 3–17, 2020. MMWR Morb Mortal Wkly Rep 2021; 70. Available at: https://www.cdc.gov/mmwr/volumes/70/wr/mm7003e3.htm. Accessed 4 August 2021.

40. Pollock NR, Jacobs JR, Tran K, et al. Performance and Implementation Evaluation of the Abbott BinaxNOW Rapid Antigen Test in a High-Throughput Drive-Through Community Testing Site in Massachusetts. J Clin Microbiol 2021; 59:e00083–21.

