## Supplement for "Testing Frequency Matters | An Evaluation of the Diagnostic Performance of a SARS-CoV-2 Rapid Antigen Test in United States Correctional Facilities"

**Rapid Antigen Test Series Definition and Test Accuracy Samples:** We defined rapid antigen test series as rapid antigen tests collected within one and four days of each other or tests collected in the absence of any test in the prior or following four days. Series were matched to RT-PCRs collected within one day prior to or following the series. If more than one RT-PCR matched to a series, we preferentially selected positive RT-PCRs followed by those collected on the same day or between rapid antigen tests, and RT-PCRs collected prior to the series.

*eFigure 1: Depiction of rapid antigen test series and the selected matched RT-PCR*

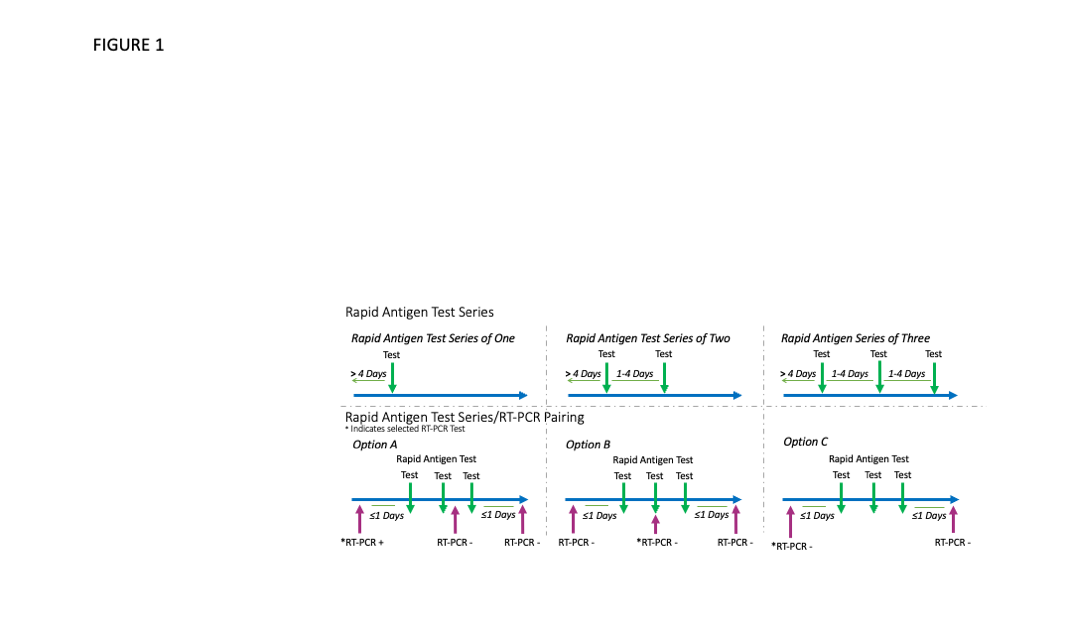

**Serial Diagnostic Accuracy Estimates:**

*eFigure 2: Serial test collection accuracy estimation approach*

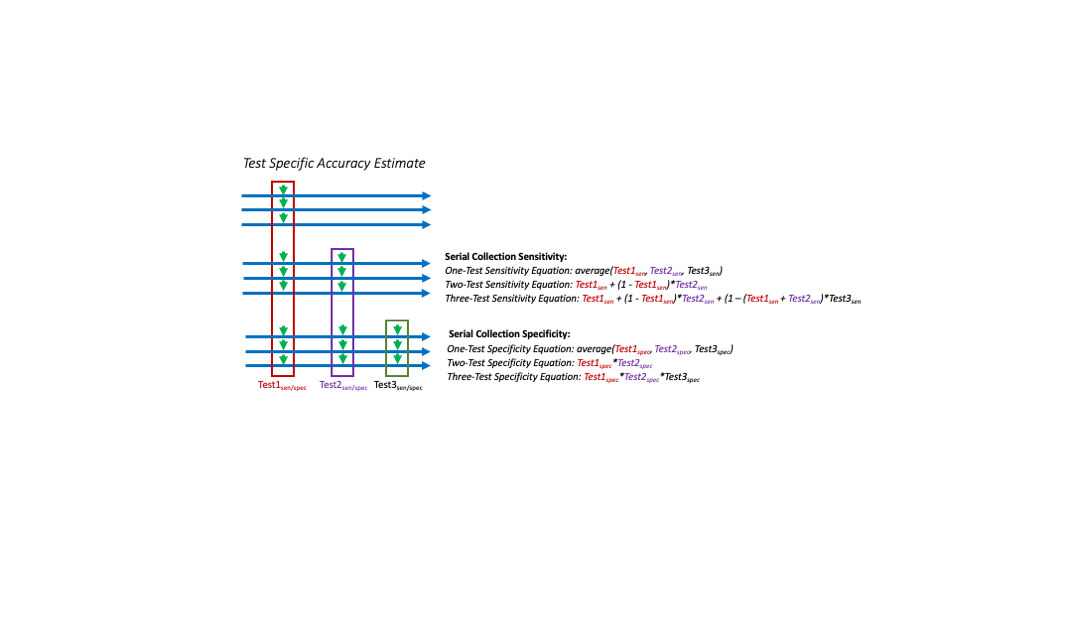

**Paired Test Series Alignment**

*eTable 1: Assay Results by Pair Type*

|  | **First Rapid Antigen Test** | **Second Rapid Antigen Test** | **Third Rapid Antigen Test** |
| --- | --- | --- | --- |
|  | **(N=3884)** | **(N=692)** | **(N=358)** |
| Concordant Results |  |  |  |
| Neither Positive | 3274 (84.3%) | 511 (73.8%) | 288 (80.4%) |
| Both Positive | 304 (7.8%) | 84 (12.1%) | 40 (11.2%) |
| Discordant Results |  |  |  |
| Rapid Antigen Test Positive | 21 (0.5%) | 8 (1.2%) | 1 (0.3%) |
| RT-PCR Positive | 285 (7.3%) | 89 (12.9%) | 29 (8.1%) |

**Testing Characteristics of RT-PCR Paired Rapid Antigen Test Series**

| *eTable 2: Testing Characteristics for Each RT-PCR Paired Rapid Antigen Test Series* | | | | | | |
| --- | --- | --- | --- | --- | --- | --- |
|  | **First Rapid Antigen Test** | | **Second Rapid Antigen Test** | | **Third Rapid Antigen Test** | |
|  | **(N=3884)** | | **(N= 692)** | | **(N=358)** | |
| Test Order |  |  |  |  |  |  |
| Rapid Antigen Test After | 696 | 17.9% | 109 | 15.8% | 68 | 19.0% |
| Rapid Antigen Test Before | 1447 | 37.3% | 430 | 62.1% | 85 | 23.7% |
| Same day/Within Set | 1741 | 44.8% | 153 | 22.1% | 205 | 57.3% |
| Symptom Presence^a^ |  |  |  |  |  |  |
| Asymptomatic | 3594 | 92.5% | 620 | 89.6% | 329 | 91.9% |
| Symptomatic | 281 | 7.2% | 72 | 10.4% | 29 | 8.1% |
| ^a^ Presence or absence of symptoms reported at the time of the rapid antigen test collection | | | | | | |

| **Predictive Value of Rapid Antigen Test Relative to RT-PCR**  *eTable 3: Rapid Antigen Test Positive and Negative Predictive Value Relative to RT-PCR Among Residents of Connecticut State Correctional Facilities Under Varying Collection Strategies* | | | | | |
| --- | --- | --- | --- | --- | --- |
|  | **Positive Predictive Value (95% CI)^c^** | | | **Negative Predictive Value (95% CI)^c^** | |
|  | **Single Test^a,c^** | **Testing Strategy^b,c^** | **Single Test^a,c^** | | **Testing Strategy^b,c^** |
| First Rapid Antigen Test | 70.8 (61.4, 78.7%) | 63.5 (49.4, 74.6%) | 98.6 (98.4, 98.7%) | | 98.6 (98.4, 98.7%) |
| Second Rapid Antigen Test | 49.0 (32.3, 65.6%) | 50.9 (38.5, 62.5%) | 98.5 (98.2, 98.7%) | | 99.2 (99.1, 99.4%) |
| Third Rapid Antigen Test | 79.3 (41.8, 97.5%) | 50.1 (35.6, 62.5%) | 98.8 (98.4, 99.1%) | | 99.7 (99.6, 99.8%) |
| ^a^ 95% confidence intervals estimated using generalized estimating equations with robust standard errors when >1 test pair per person was present, alternatively, Wald confidence intervals were estimated | | | | | |
| ^b^ Serial testing sensitivity was estimated as the additive probability (positive for any rapid antigen test); serial testing specificity was estimated as the multiplicative probability (negative for all rapid antigen test), posterior simulation of 1000 draws was used to propagate uncertainty through the equations | | | | | |
| ^c^ Positive predictive value (PPV) and negative predictive value (NPV) estimated using the estimated prevalence, sensitivity, and specificity | | | | | |

**Sensitivity Analyses:** To conduct the presented analysis, we had to make decisions regarding which tests to include. Because the selected tests may not reflect the exact timing of tests collected in practice, this selection process may have introduced bias into our analysis. In the following sensitivity analyses, we estimate the diagnostic accuracy under varying test selection scenarios. Outside of the specific scenario described in the sensitivity analysis, the approach and sample matched that of the primary analysis.

*Exact three days between tests:* While the CT DOC recommends rapid antigen tests be collected serially every 3 days for up to 3 negative tests, test collection within facilities is difficult and adhering to this strict testing schedule may not always be feasible. For this reason, we included the tests collected within 1 and 4 days of each other in our rapid antigen test series. To test if this loose definition of serial testing drove our results, we performed a sensitivity analysis where we restricted to serial tests collected exactly three days apart.

| *eTable 4: BinaxNOW Accuracy Relative to RT-PCR Among Residents of Connecticut State Correctional Facilities Under Varying Testing Strategies (Exactly three days)* | | | | | | |
| --- | --- | --- | --- | --- | --- | --- |
|  | **No. RT-PCR Positive Pairs** | **Sensitivity (95% CI)** | | **No. RT-PCR Negative Pairs** | **Specificity (95% CI)** | |
| **Characteristics** |  | **Single Test^a^** | **Testing Strategy^b^** |  | **Single Test^a^** | **Testing Strategy^b^** |
| First Rapid Antigen Test | 546 | 63.6 (59.4, 67.5%) | 75.4 (62.7, 88.0%) | 3474 | 99.3 (99.0, 99.6%) | 99.61 (98.33, 100%) |
| Second Rapid Antigen Test | 76 | 61.8 (50.7, 72.2%) | 86.1 (81.8, 89.9%) | 326 | 99.7 (98.7, 100%) | 98.86 (97.3, 99.4%) |
| Third Rapid Antigen Test | 8 | 100 (63.1, 100%) | 100 (95.0, 100%) | 97 | 100 (96.3, 100%) | 98.85 (95.0, 100%) |
| ^a^ 95% confidence intervals estimated using Generalized Estimating Equations with robust standard errors when >1 test pair per person was present, alternatively, Wald confidence intervals were estimated | | | | | | |
| ^b^ Serial testing sensitivity was estimated as the additive probability (positive for any rapid antigen test); serial testing specificity was estimated as the multiplicative probability (negative for all rapid antigen tests); posterior simulation of 1000 draws was used to propagate uncertainty through the equations | | | | | | |

*Random selection of rapid antigen tests:* In our primary analysis, we assumed that the observed first, second and third rapid antigen tests were representative of the tests under perfect serial collection (all residents were tested three times if all tests were negative). However, such collection is challenging and 2,869 of negative RT-PCR matched rapid antigen tests were collected in isolation. As a result, the second and third rapid antigen samples may be biased. An alternative approach is to randomly select which tests end up in the first, second, and third position of each series. The following are the results from such an analysis. For this analysis, we kept the number of tests to be the same as in the primary analysis (3884 first tests, 692 second tests, and 358 third tests). To ensure we account for the randomness of our selection, we provided estimates from four iterations in table form and graphed results from 1000 iterations.

*eFigure 3: Sensitivity and specificity of seriall collection strategies for randomly selected samples*

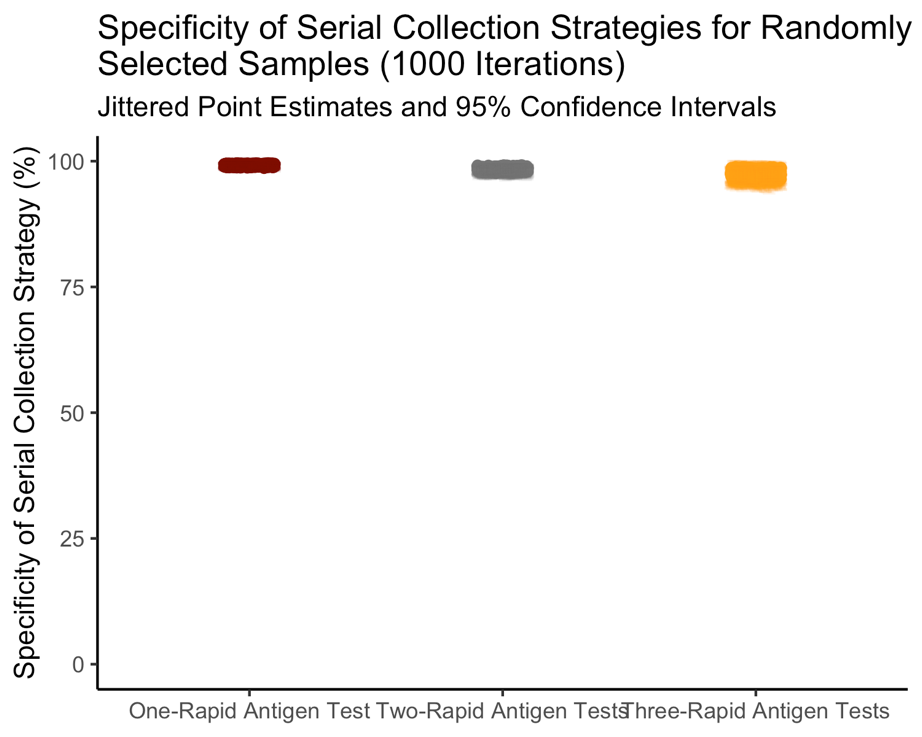

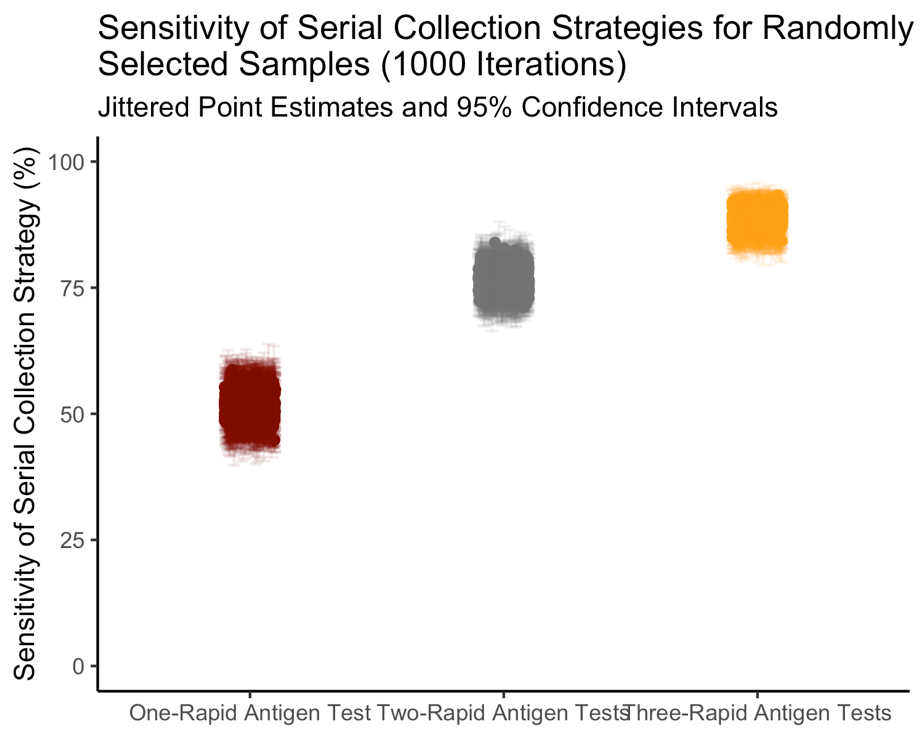

| *eTable 5: Rapid Antigen Test Accuracy Relative to RT-PCR Among Residents of Connecticut State Correctional Facilities Under Varying Collection Strategies (Random test selection)* | | | | | | |
| --- | --- | --- | --- | --- | --- | --- |
|  | **No. RT-PCR Positive Pairs** | **Sensitivity (95% CI)** | | **No. RT-PCR Negative Pairs** | **Specificity (95% CI)** | |
|  |  | **Single Test^a^** | **Testing Strategy^b^** |  | **Single Test^a^** | **Testing Strategy^b^** |
| *Iteration 1* |  |  |  |  |  |  |
| First Rapid Antigen Test | 647 | 51.0 (47.2, 54.8%) | 52.4 (47.2, 57.6%) | 3237 | 99.3 (98.9, 99.5%) | 99.3 (98.6, 99.6%) |
| Second Rapid Antigen Test | 122 | 54.1 (45.3, 62.7%) | 77.6 (73.0, 81.9%) | 571 | 99.1 (98.1, 99.7%) | 98.3 (97.2, 98.9%) |
| Third Rapid Antigen Test | 62 | 51.6 (39.3, 63.8%) | 89.2 (85.4, 92.6%) | 296 | 99.7 (98.5, 100%) | 97.8 (95.9, 98.7%) |
| *Iteration 2* |  |  |  |  |  |  |
| First Rapid Antigen Test | 652 | 51.5 (47.7, 55.4%) | 51.8 (46.6, 57.2%) | 3232 | 99.3 (99.0, 99.6%) | 99.0 (98.3, 99.5%) |
| Second Rapid Antigen Test | 124 | 50.8 (42.1, 59.5%) | 76.3 (71.7, 80.6%) | 569 | 99.3 (98.4, 99.8%) | 98.5 (97.4, 99.1%) |
| Third Rapid Antigen Test | 55 | 52.7 (39.6, 65.4%) | 88.8 (84.9, 92.3%) | 303 | 98.7 (97.0, 99.6%) | 97.1 (95.0, 98.4%) |
| *Iteration 3* |  |  |  |  |  |  |
| First Rapid Antigen Test | 648 | 52.3 (48.5, 56.1%) | 49.5 (44.4, 54.8%) | 3236 | 99.4 (99.0, 99.6%) | 99.1 (98.5, 99.5%) |
| Second Rapid Antigen Test | 121 | 51.2 (42.4, 60.0%) | 76.8 (72.3, 81.1%) | 572 | 99.0 (97.9, 99.6%) | 98.2 (97.0, 98.9%) |
| Third Rapid Antigen Test | 63 | 44.4 (32.7, 56.8%) | 87.2 (83.3, 90.9%) | 295 | 99.3 (97.9, 99.9%) | 97.4 (95.5, 98.5%) |
| *Iteration 4* |  |  |  |  |  |  |
| First Rapid Antigen Test | 627 | 52.6 (48.7, 56.5%) | 49.5 (44.4, 54.8%) | 3257 | 99.3 (99.0, 99.6%) | 99.0 (98.3, 99.5%) |
| Second Rapid Antigen Test | 139 | 46.8 (38.6, 55.1%) | 76.8 (72.3, 81.1%) | 554 | 99.6 (98.9, 99.9%) | 98.9 (97.9, 99.3%) |
| Third Rapid Antigen Test | 66 | 51.5 (39.6, 63.3%) | 87.2 (83.2, 90.9%) | 292 | 98.3 (96.4, 99.4%) | 97.0 (94.9, 98.4%) |
| ^a^ 95% confidence intervals estimated using Generalized Estimating Equations with robust standard errors when >1 test pair per person was present, alternatively, Wald confidence intervals were estimated | | | | | | |
| ^b^ Serial testing sensitivity was estimated as the additive probability (positive for any rapid antigen test); serial testing specificity was estimated as the multiplicative probability (negative for all rapid antigen tests); posterior simulation of 1000 draws was used to propagate uncertainty through the equations | | | | | | |

*Ordered selection if multiple RT-PCR matches:* In the event of more than one RT-PCR matched with a rapid antigen series, we had to select down to one. In the primary analysis, we gave ordered preference based on positivity, same day or between rapid antigen collection, and RT-PCR collected before the rapid antigen. To ensure this selection process did not drive our results, we altered the ordered selection in two ways:

1. Negative RT-PCR, same day collection (or between rapid antigen tests), RT-PCR collected before rapid antigen
2. Same day of collection (or between rapid antigen tests), RT-PCR collected before rapid antigen test, Positive RT-PCR

| *eTable 6: BinaxNOW Accuracy Relative to RT-PCR Among Residents of Connecticut State Correctional Facilities Under Varying Collection Strategies (Varying RT-PCR selection in the event of multiple matches)* | | | | | | |
| --- | --- | --- | --- | --- | --- | --- |
|  | **No. RT-PCR Positive Pairs** | **Sensitivity (95% CI)** | | **No. RT-PCR Negative Pairs** | **Specificity (95% CI)** | |
| **Characteristics** |  | **Single Test^a^** | **Testing Strategy^b^** |  | **Single Test^a^** | **Testing Strategy^b^** |
| *Negative RT-PCR, same day collection (or between rapid antigen tests), RT-PCR collected before rapid antigen test* | | | | | |  |
| First Rapid Antigen Test | 585 | 51.8 (47.7, 55.8%) | 52.8 (48.1, 57.7%) | 3299 | 99.3 (99.0, 99.6%) | 99.0 (98.2, 99.4%) |
| Second Rapid Antigen Test | 171 | 49.1 (41.7, 56.6%) | 75.6 (71.4, 79.5%) | 521 | 98.5 (97.2, 99.3%) | 97.7 (96.3, 98.6%) |
| Third Rapid Antigen Test | 68 | 57.4 (45.5, 68.7%) | 89.6 (86.1, 92.7%) | 290 | 99.3 (97.9, 99.9%) | 96.9 (94.8, 98.1%) |
| *Same collection day* (or between rapid antigen tests)*, RT-PCR collected before rapid antigen test, positive RT-PCR* | | | | | | |
| First Rapid Antigen Test | 589 | 51.6 (47.6, 55.6%) | 52.8 (48.1, 57.6%) | 3295 | 99.4 (99.1, 99.6%) | 99.1 (98.4, 99.5%) |
| Second Rapid Antigen Test | 173 | 48.6 (41.2, 56.0%) | 75.2 (71.0, 79.2%) | 519 | 98.5 (97.2, 99.3%) | 97.8 (96.4, 98.6%) |
| Third Rapid Antigen Test | 69 | 58.0 (46.2, 69.2%) | 89.6 (86.1, 92.6%) | 289 | 99.7 (98.5, 100%) | 97.2 (95.1, 98.5%) |
| ^a^ 95% confidence intervals estimated using Generalized Estimating Equations with robust standard errors when >1 test pair per person was present, alternatively, Wald confidence intervals were estimated | | | | | | |
| ^b^ Serial testing sensitivity was estimated as the additive probability (positive for any rapid antigen test); serial testing specificity was estimated as the multiplicative probability (negative for all rapid antigen tests); posterior simulation of 1000 draws was used to propagate uncertainty through the equations | | | | | | |
